# Knowledge, Attitudes and Demand Toward Cardiovascular Polygenic Risk Testing in Clinical Practice: Cross-Sectional Survey of Patients

**DOI:** 10.1101/2023.08.24.23294594

**Authors:** Shanjot Brar, Jared Townsend, Joban Phulka, Laura Halperin, Janet Liew, Jeremy Parker, Liam R. Brunham, Zachary Laksman

**Author notes:** Corresponding authors: a. Shanjot Brar, MD Gordon & Leslie Diamond Health Care Centre 2775 Laurel St, 9th Floor, Vancouver, BC, Canada V5Z1M9. b. Zachary Laksman, MD (senior corresponding author) 1033 Davie St Suite 211, Vancouver, BC V6E 1M7.

## Abstract

**Background:** The goal of this study was to assess patients’ prior exposure and current level of knowledge of polygenic risk scores (PRSs). We also explored reactions to receiving a high-risk or low-risk score, and gauged the overall attitudes and demand patients have with regards to PRSs.

**Methods:** We developed an online investigator-designed survey based on existing validated tools and previously designed surveys on genetic testing. There were two versions of the survey, one including a hypothetical high-risk PRS and one with a low-risk PRS. We administered the survey among patients attending a specialized cardiovascular prevention clinic.

**Results:** A total of 226 participants responded to the survey. The study population was predominantly high-income earning, educated, and of European descent. 177 patients (79%) had never read or heard about polygenic testing. 209 patients (93%) had never discussed polygenic testing with their health care professional (HCP). 208 patients (93%) had never received polygenic testing.

The average score on the knowledge quiz was 2.47/10 [95% C.I. (2.17, 2.78)]. Participants that received a high-risk survey scored 20.52/35 [95% C.I. (16.14, 24.9)] with regards to negative emotions while low-risk survey participants scored 17.96/35 [95% C.I. (13.98, 21.94)] (p<0.001). Participants that received a high-risk survey scored 5.78/10 [95% C.I. (3.77, 7.79)] with regards to uncertainty and low-risk survey participants scored 4.34/10 [95% C.I. (2.50, 6.18)] (p<0.001). Participants that received a high-risk survey scored 12.42/15 [95% C.I. (10.43, 14.41)] for demand and low-risk survey participants scored 12.22/15 [95% C.I. (9.66, 14.78)] (p=0.549).

**Conclusions:** Patients have limited prior exposure and knowledge of PRSs. Compared to receiving a low-risk score, participants receiving a high-risk score have more negative emotions and feelings of uncertainty. Despite the lack of knowledge, and the high rate of negative emotions and uncertainty, demand for PRSs in cardiology practice is high and expected to increase.

## INTRODUCTION

Polygenic risk scores (PRSs) are being increasingly used to help predict diseases with complex inheritance patterns that have thus far not been explained by mendelian inheritance, such as atrial fibrillation and coronary artery disease. ^1,2^ PRSs also have the potential to contribute to the developing field of personalized medicine beyond risk prediction, and may inform personalized treatment strategies.^3^ A recent American Heart Association (AHA) scientific statement explored the science, clinical considerations and future challenges of PRSs for cardiovascular disease and concluded that the addition of PRSs to clinical risk tools consistently enhances predictive ability.^4^ There are many technical limitations to PRSs, however beyond this, the current workforce is not equipped to utilize PRSs in clinical practice due to insufficient knowledge, training, and tools.^5^ Patients and consumers have minimal exposure and experience incorporating complex genomic information and probabilities into their decision making.^6^ Studies have investigated patient responses to PRSs in other clinical diseases^1,7,8^, however such information is lacking in the cardiovascular space. Understanding these care gaps will help to inform future implementation strategies.

In this study, we directly tested the hypothesis that patients are poorly prepared to receive and integrate genetic results, including cardiovascular PRSs. To do this, we assessed patients’ prior exposure and current level of knowledge of PRSs. We also explored patients’ reactions to receiving a high-risk or low-risk result. Finally, we gauged the overall attitudes and demand patients have with regards to PRSs.

## METHODS

We used an online survey with quantitative responses through the University of British Columbia (UBC) Survey tool (Supplemental Appendix). The UBC Survey tool is a Canadian-hosted program that is compliant with the British Columbia Freedom of Information and Protection Act. This project was approved by the UBC Research Ethics Board, REB# H22-02087. Initially, a consecutive chart review of patients that were attending at the St. Paul’s Hospital Healthy Heart Program was conducted. The Healthy Heart Program is a quaternary referral centre located in Vancouver, British Columbia, which focuses on primary and secondary cardiovascular disease prevention. Patients that had previously consented to be contacted for research were identified from chart review. These patients were then individually contacted via phone call to consent to receive an email to participate. Patients who provided verbal consent were then individually sent an email which included a consent form and a link to the survey. Participants were randomly assigned 1:1 to receive either a high-risk or low-risk version of the survey. An example recruitment email is provided in the supplemental appendix.

The survey design was based on existing validated tools and previously designed surveys on genetic testing^1,7–13^ and was divided into the following components: a) Demographics, b) Prior knowledge, c) Knowledge of polygenic risk scores, d) Educational video e) Response toward a “high-risk” or “low-risk score” and f) Demand. Participant knowledge was assessed using a ten-question quiz, with a total score ranging from 0-10. The high-risk survey included an example of a polygenic risk score percentile of >95%, while the low-risk survey included an example of a polygenic risk score percentile <5%. The Feelings About genomiC Testing Results (FACToR) scale was adapted and used to assess participant reactions to high-risk and low-risk results.^1^ Negative emotions were assessed using seven statements, with each statement being scored from a range of 0-5 (1 = Not at all, 5 = A great deal). Uncertainty was assessed using two statements with each statement being scored from a range of 1-5 (1 = Not at all, 5 = A great deal). A total score was then tabulated for both negative emotions (minimum = 7, maximum = 35) and uncertainty (minimum = 2, maximum = 10). A higher score indicated higher uncertainty, and more negative emotions. Participant demand for PRSs was assessed using three separate statements with each statement being scored from a range of 0-5 (1 = strongly disagree, 5 = strongly agree). A total score was then tabulated (minimum = 3, maximum = 15). A higher score indicated greater demand. The survey in its entirety, including both high-risk and low-risk examples, as well as the educational video are provided in the supplemental material.

## RESULTS

A total of 1,756 patient charts were reviewed, of which 1,130 patients who had expressed interest in research participation were identified. These patients were individually contacted via telephone and 366 patients consented to participate in our survey. Half the participants were sent the high-risk survey (n=183) and half were sent the low-risk survey (n=183) via email. 118 participants responded to the high-risk survey and 108 participants responded to the low-risk survey (Figure 1).

**Figure 1.**
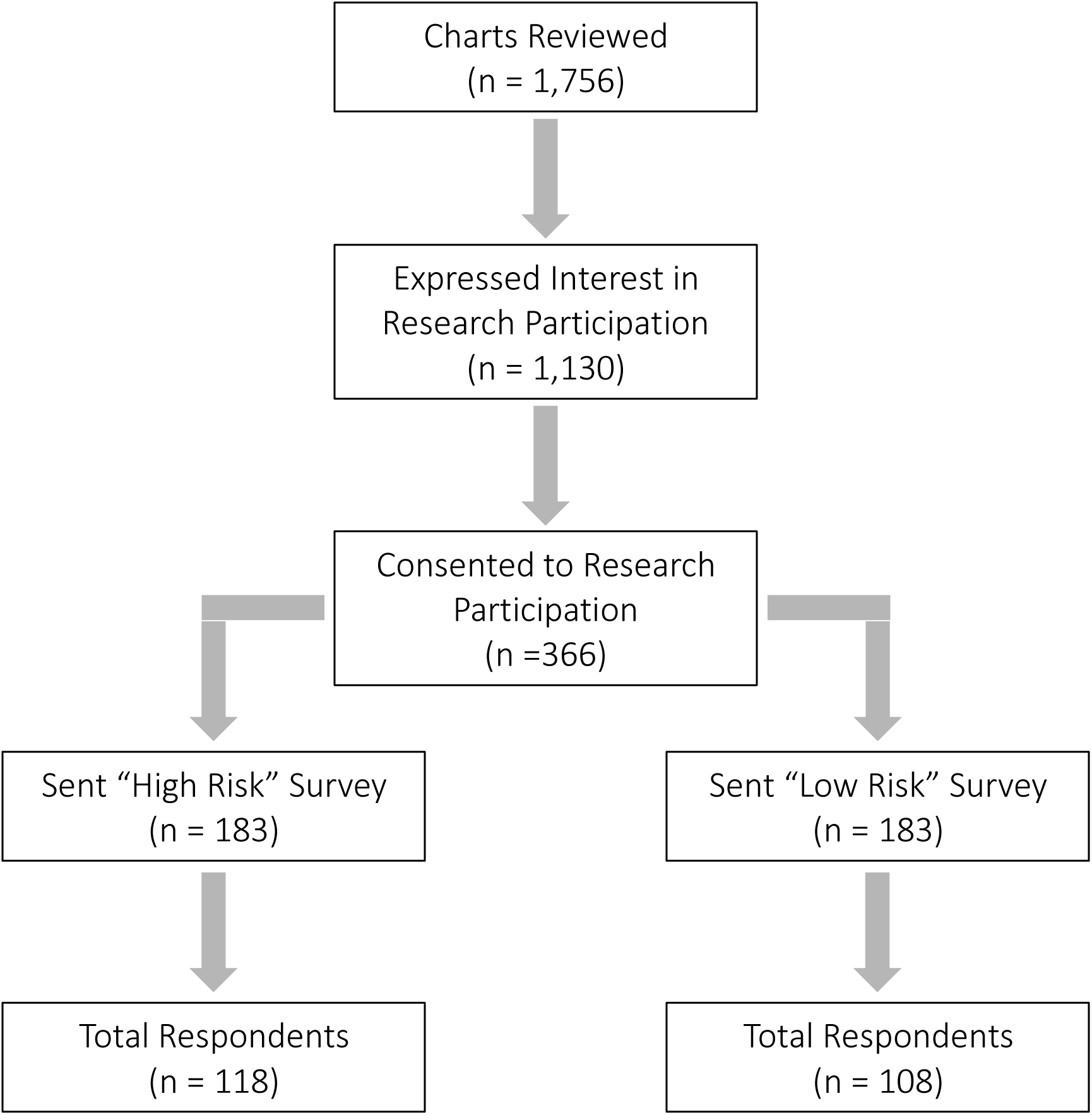
Study enrollment.

Participant characteristics including age, gender, highest level of education, household income and ethnicity are listed in Table 1. 177 patients (79%) had never read or heard about polygenic risk scores. 209 patients (93%) had never discussed polygenic risk scores with their health care professional (HCP). 208 patients (93%) had never received polygenic risk score results (Figure 2). The average score on the knowledge quiz was 2.47/10 [95% C.I. (2.17, 2.78)].

**Figure 2.**
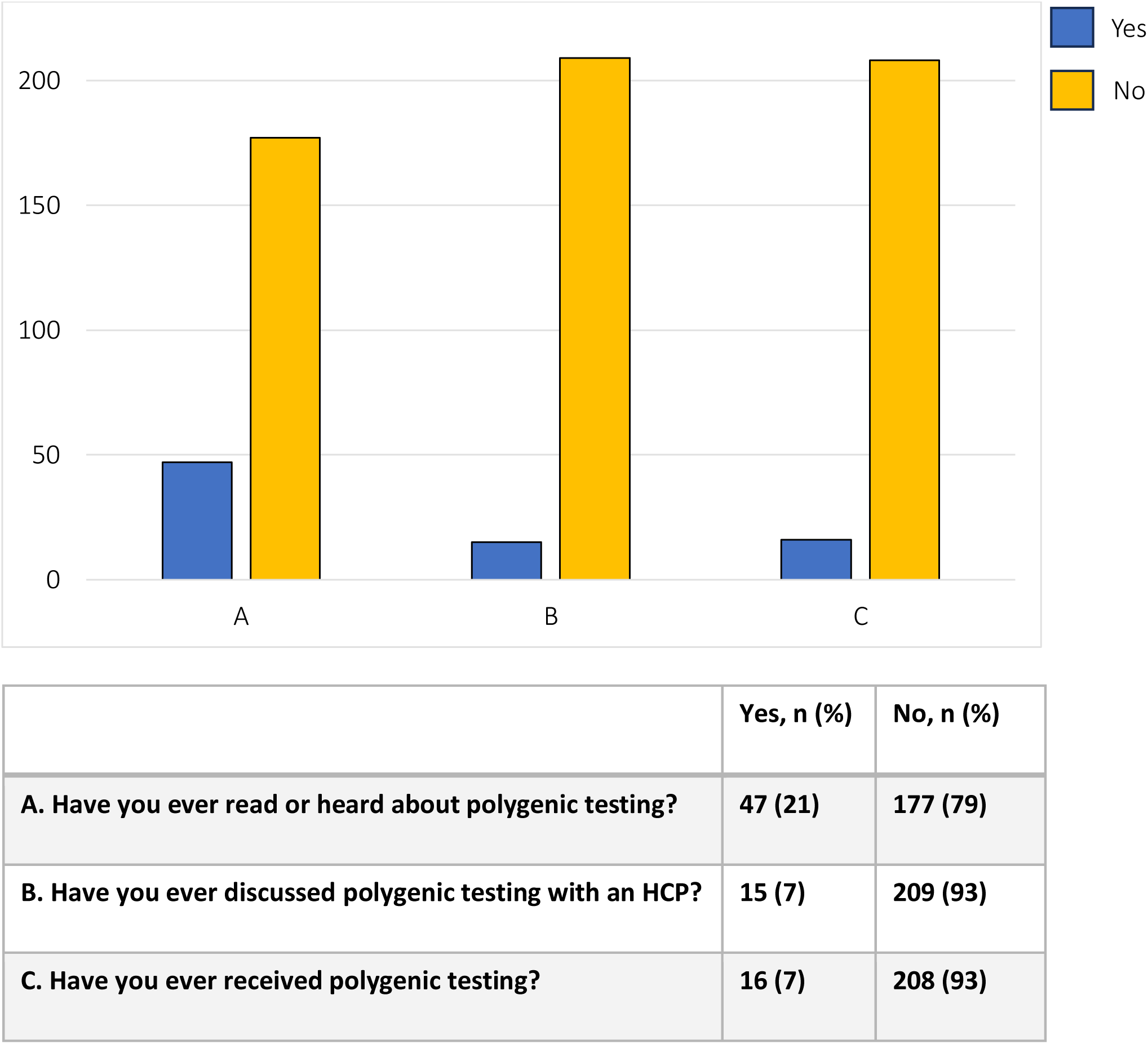
Participant prior knowledge of Polygenic Risk Scores (PRS)

**Table 1.** Patient demographics.

|  |  |
| --- | --- |
| Age, n (%) |  |
| 18-30 years | 5 (2) |
| 31-40 years | 10 (4) |
| 41-50 years | 27 (12) |
| 51-60 years | 57 (25) |
| >60 years | 127 (56) |
| Gender, n (%) |  |
| Male | 121 (54) |
| Female | 105 (46) |
| Highest level of education, n (%) |  |
| Did not Finish High School | 6 (3) |
| High School | 27 (12) |
| Post-Secondary | 77 (34) |
| Bachelor’s Degree | 77 (34) |
| Master’s Degree | 21 (9) |
| Doctorate/PhD | 18 (8) |
| Household income, n (%) |  |
| <\$50,000 | 37 (16) |
| \$50,000 – \$150,000 | 110 (49) |
| >\$150,000 | 79 (35) |
| Ethnicity, n (%) |  |
| African | 1 (0) |
| European | 155 (69) |
| East Asian | 15 (7) |
| South Asian | 11 (5) |
| Southeast Asian | 6 (3) |
| First Nations/Indigenous | 2 (1) |
| Hispanic | 1 (0) |
| Middle Eastern | 2 (1) |
| Other | 33 (15) |

Participants that received a high-risk survey scored 20.52/35 [95% C.I. (16.14, 24.9)] with regards to negative emotions while participants that received a low-risk survey scored 17.96/35 [95% C.I. (13.98, 21.94)] (p<0.001). Participants that received a high-risk survey scored 5.78/10 [95% C.I. (3.77, 7.79)] with regards to uncertainty while participants that received a low-risk survey scored 4.34/10 [95% C.I. (2.50, 6.18)] (p<0.001) (Table 2).

**Table 2.** Participant negative emotions and uncertainty scores for polygenic risk scores.

|  | High Risk | Low Risk | P value |
| --- | --- | --- | --- |
|  | N = 118 | N = 108 |  |
| Negative Emotion |  |  |  |
| Total Score (/35) ± SD | 20.52 ± 4.38 | 17.96 ± 3.98 | <0.001 |
| Uncertainty |  |  |  |
| Total Score (/10) ± SD | 5.78 ± 2.01 | 4.34 ± 1.84 | <0.001 |

Participant demand for PRS testing was high overall 12.32/15 [95% C.I. (11.99, 12.65)] and was not significantly different between individuals who received a high-risk survey 12.42/15 [95% C.I. (10.43, 14.41)] compared to a low-risk survey 12.22/15 [95% C.I. (9.66, 14.78)] (p=0.549) (Table 3).

**Table 3.** Participant demand for polygenic risk scores.

|  | High Risk | Low Risk | P value |
| --- | --- | --- | --- |
|  | N = 118 | N = 108 |  |
| Belief |  |  |  |
| Total Score (/15) ± SD | 12.42 ± 1.99 | 12.22 ± 2.56 | 0.549 |

## DISCUSSION

PRSs have a promising future application for identifying individuals at risk of cardiovascular disease and creating more individualized treatment plans, however, there are many technical and systematic limitations that must be overcome prior to their inclusion in routine cardiovascular care.^2,4^ As these limitations are overcome, and PRSs are used more regularly in routine care, it is increasingly important that we understand patient perspectives as we train our future workforce and develop implementation strategies across different health systems. Previous studies have investigated patient responses to PRSs in other clinical diseases ^1,7,8^, however to our knowledge, this is the first study examining patient perspectives in the cardiovascular space.

One of the proposed goals of PRSs in clinical practice is to use genetic based risk as a means of promoting healthy behaviours and motivating high risk individuals to make lifestyle changes.^4,6,15,16^ Numerous studies however have failed to show a change in patient behaviour by communicating genetic risk.^17–19^ Knowles et. al. demonstrated no major effect of communicating a genetic risk score for CAD in reducing certain health risk behaviours.^20^ Conversely, studies have shown behaviour modification associated with patients who underwent coronary artery calcium (CAC) testing.^21,22^ Although patients’ behavioural response to genetic risk is influenced by multiple factors, a significant barrier includes patients’ understanding and interpretation of genetic risk. ^16,23,24^ Many authors have suggested that patients’ understanding of cardiovascular PRSs is poor.^25^ Our study confirms this finding and despite the education level of our surveyed patients, 177 patients (79%) had never read or heard about polygenic testing, and the average score on the knowledge quiz was 2.47/10 (24.7%). Moreover, the survey population was selected from a group of patients already followed at a sub-specialized cardiovascular disease prevention clinic, and an even lower level of exposure and knowledge would be expected if this survey was offered to a broader and more diverse population.

Genetics related literacy impacts the value patients place on genetic information as well as how patients evaluate the utility of genetic testing.^13,26^ Low genetic literacy has also been associated with poor understanding of the limitations of genetic testing.^27^ This implies that a low knowledge of genetic testing, as seen in our study, would result in a significant risk of misinterpretation or misapplication of genetic risk scoring. Although this phenomenon has not been explicitly reported in the cardiovascular space, it is well documented in other clinical fields that integrate genetic testing.^12,27^ It is unclear, the level of knowledge or the methods of knowledge translation that are required for the clinical implementation of cardiovascular PRSs, and further understanding of this topic is needed. Furthermore, it has already been identified both by the CDC and the NIH that the current workforce of clinicians and health care providers are undertrained and poorly suited to provide the requisite education regarding complex probabilistic polygenic testing.^28,29^

Our results also demonstrate that patients given a high-risk PRS report higher levels of negative emotions and uncertainty about PRSs than those who received a low-risk PRS. This was an expected finding, as previous studies have shown, that patients will often perceive health data as threats and the recommended behavioural change will be quite different from their health care provider.^23^ This finding is seen even more commonly when it is related to genetic information.^30,31^ This demonstrates a dilemma for clinicians and health care providers as patients with high-risk PRS will likely require more intensive risk modifying treatment plans and closer monitoring but may also have a significant amount of uncertainty and negative emotions to overcome. Previous studies have shown that patient interpretation of genetic risk information is not only related to the statistical findings, but also to patients’ uncertainties related to the topic.^32,33^ The higher degree of uncertainty and negative emotions will be an obstacle for clinical implementation and must be accounted for in future integration of PRSs.

Despite the high level of negative emotions and uncertainty, surveyed patients reported that the health benefits of PRS outweigh the risks (79.6% responded agree or strongly agree) and believe that polygenic testing should be included in heart disease prevention programs for the general public (81.9% responded agree or strongly agree). Furthermore, surveyed patients reported that they would like PRS included in their care plans (85.3% responded agree or strongly agree). Taken together, this suggests a high demand for use of PRSs in clinical practice. This trend was demonstrated across all participant groups, regardless of whether they received a high-risk score, or a low-risk score. These findings were expected, as a demand for genetic testing in the general population is high, as exemplified by the estimated 26 million people that had used online direct-to-consumer (DTC) genetic testing by the end of 2018.^15^ A high demand for PRSs is a promising finding for the future implementation of PRSs, however as highlighted above, the majority of patients have poor knowledge and understanding of PRSs, and thus the potential for misapplication remains quite high.

The future of PRSs includes a potential for individualized screening, preventative measures, and pharmacotherapy. This brings a slew of challenges for clinicians and health care providers, including how to determine the best ways to communicate the science. Guidelines on how to navigate communication of genetic information to patients with cardiovascular disease such as inherited rhythm disorders, hypertrophic cardiomyopathy and familial hypercholesteremia exist, however such guidelines are lacking for PRSs.^34,35^ Importantly clinicians must be able to effectively communicate the benefits, risk and limitations of PRSs.^6^ There is clinical need to develop educational materials for both patients and guidelines for clinicians on how to convey this information. Knowledge translation in PRSs will be a great challenge moving forward, however there are many evidence-based strategies that exists for the presentation of genetic risk.^36,37^ Incorporating these lessons to PRSs will be pivotal to ensure patients understand and can effectively engage in their care. Lessons can also be learned from shared decision making tools in the cardiovascular space, such as tools to help guide patient centered conversations about anticoagulation in atrial fibrillation.^38^ Previous studies have shown that such shared decision making tools can significantly lower decisional conflict between patients and clinicians.^39^ Development of similar tools for PRSs would potentially enhance patient and physician experience with navigating discussion and implementation of PRSs.

## Limitations

Our study has important limitations, largely related to the selection of our patient population. The study population was predominantly high-income earning, educated, and of European descent. As our survey required consent at two different stages prior to distribution of the survey, this specific population is likely a result of selection bias and may indicate that this specific population is more interested in PRSs. This would need further investigation, however would contribute to the growing concern regarding widening care disparities related to PRSs.^14^ Further, our method of accessing patients was through a specialized cardiovascular disease prevention clinic where patient attendance likely indicates their want, or need, to make risk factor modifications. This would result in significant selection bias and not necessarily represent a routine cardiology clinic. Additionally, we accessed patients that had already consented to research involvement due to the process of ethical approval. As such these patients are likely highly engaged in their care and may not represent the patient population in contemporary clinical practice.

## Conclusions

Patients attending a specialized cardiovascular clinic focused on primary and secondary prevention had limited prior exposure and knowledge with regards to PRSs. When compared to receiving a low-risk score, participants that receive a high-risk score have more negative emotions and feelings of uncertainty. Despite the lack of knowledge, and the high rate of negative emotions and uncertainty, patient demand for PRSs in contemporary cardiology practice is high. The analytical aspects of PRSs are continually being addressed, however there needs to be concurrent focus on improving patient and provider exposure and knowledge to avoid future harm. Future studies should focus on the development of educational materials and guidelines to address the barriers brought forward by this study.

## DISCLOSURES

The authors have no conflicts to disclose.

### ABBREVIATIONS

Non-standard Abbreviations and Acronyms:

PRS(s): polygenic score(s)
AHA: American Heart Association
UBC: University of British Columbia
FACToR: Feelings About genomiC Testing Results
CAD: coronary artery disease
CAC: coronary artery calcium
CDC: Centres for Disease Control and Prevention
NIH: National Institutes of Health
DTC: direct to consumer

## Data Availability

The data that support the findings of this study are available on request from the corresponding author

## SUPPLEMENTAL MATERIAL

Material attached separately.

